# Video-based gait analysis using pose estimation can quantify gait differences among non-frail, pre-frail, and frail older adults

**DOI:** 10.64898/2026.08.04.26359742

**Authors:** Kaleb Burch, Jaya Hamkins, Laura McDaniel, Ana Raquel Castro e Costa, Zechen Yang, Jan Stenum, Andrew Pagliocchini, Crystal Szczesny, Jacqueline Langdon, Rama Chellappa, Peter M. Abadir, Ryan T. Roemmich

**Affiliations:** Department of Medicine, Division of Geriatric Medicine and Gerontology, The Johns Hopkins University School of Medicine, Baltimore, MD 21224; Center for Movement Studies, The Kennedy Krieger Institute, Baltimore, MD 21205; Department of Biomedical Engineering, The Johns Hopkins University School of Medicine, Baltimore, MD 21205; Department of Electrical and Computer Engineering, The Johns Hopkins University, Baltimore, MD 21218; Department of Neuroscience, The Johns Hopkins University School of Medicine, Baltimore, MD 21205; Department of Physical Medicine and Rehabilitation, The Johns Hopkins University School of Medicine, Baltimore, MD 21205

**Keywords:** frailty, aging, artificial intelligence, computer vision, kinematics, walking

## Abstract

Frailty is a common consequence of aging that makes individuals increasingly susceptible to adverse health outcomes. Frailty screening can identify pre-frail and frail individuals to prescribe interventions or inform clinical decision making to prevent or slow additional frailty progression. Objective, scalable, and automated frailty assessments may expedite and improve clinical frailty screening. Here, we leveraged human pose estimation for video-based gait analysis in older adults who were non-frail, pre-frail, and frail. We focused on gait because slow walking speed is key diagnostic criteria of frailty, and many gait deviations are often observed in older adults with frailty. We collected videos of 68 older adults (25 non-frail, 25 pre-frail, 18 frail) walking at both self-selected and fast paces and used an established pose estimation-based gait analysis approach to measure and compare gait parameters across frailty statuses. Pose estimation-based step time measurements were strongly correlated with manual annotations (self-selected: R^2^=0.93, fast: R^2^=0.80) and showed tight Bland-Altman limits of agreement (self-selected: -0.082 to 0.052s, fast: -0.114 to 0.110s), establishing validity of this video-based gait analysis approach in older adults. We then identified a series of cross-sectional differences in spatiotemporal gait parameters among non-frail, pre-frail, and frail older adults, demonstrating that video-based gait analysis can be useful for measuring gait differences across frailty statuses. This study demonstrates the potential of video-based pose estimation for scalable gait tracking across frailty statuses in older adults.

## INTRODUCTION

Frailty is a common consequence of aging that is characterized by a state of increased vulnerability resulting from an age-associated decline in normal function across multiple body systems and a diminished physiological reserve (Fried et al., 2001). This vulnerability impairs the ability to cope with everyday or acute stressors and is associated with a markedly higher risk of adverse health outcomes including falls, disability, surgical complications, hospitalization, and mortality (Vermeiren et al., 2016). The recognition of frailty as a distinct medical syndrome has led to the development of standardized assessment tools that enable clinicians to identify at-risk individuals in various healthcare settings (Fried et al., 2001; Mitnitski, Mogilner, & Rockwood, 2001).

Incorporation of frailty assessments into clinical practice is known to improve risk prediction and inform clinical decision-making for older patients, especially in acute care environments (Gilbert et al., 2018). Currently, the physical frailty phenotype is the most prominent frailty assessment scale. This scale is evaluated based on five key criteria: unintentional weight loss, weakness (measured via grip strength), self-reported exhaustion, slow walking speed, and low physical activity (Fried et al., 2001). Under this assessment, all five categories are reduced to binary scores, but diagnosis and prediction of frailty could be improved with quantitative and granular assessments of motor function.

Here, we focus on gait assessment in particular because walking ability promotes independence, community engagement, and lower risk of all-cause mortality in older adults (Paluch et al., 2022). Walking speed is often considered a sixth vital sign (Middleton, Fritz, & Lusardi, 2015) and is strongly associated with falls, disability, hospitalization, and mortality (Abellan Van Kan et al., 2009; Dumurgier et al., 2009). Walking speed alone has been recommended as a single-item measure of frailty by an International Academy on Nutrition and Aging Task Force (Abellan Van Kan et al., 2009) given its reliable predictive properties on par with established assessments like the Short Physical Performance Battery (Guralnik et al., 2000). Notably, Abellan Van Kan et al., 2009 highlights the value of such an assessment because it requires patients only to walk for a short bout and because walking function depends on various physiological systems relevant to frailty.

We propose to expand on the notion of measuring frailty status with just a simple walking test by measuring various gait parameters from a single walking video. Quantitative gait analysis can expand beyond walking speed and provide data on gait kinematics (e.g., lower limb joint angles), spatiotemporal gait parameters (e.g., step length, step time), and gait variability that have important associations with aspects of mobility commonly affected by aging and frailty, including energy expenditure (Schrack, Zipunnikov, Simonsick, Studenski, & Ferrucci, 2016) and dynamic balance (Brach, Berlin, Vanswearingen, Newman, & Studenski, 2005; Gabell & Nayak, 1984; Montero-Odasso et al., 2011). Traditionally, quantitative gait analysis has been inaccessible in most clinical settings due to major barriers that include prohibitive costs, time-consuming setup and data collection procedures, and a need for technical expertise for operation. There is a clear need for new technologies that can provide quantitative, whole-body gait analyses in older adults that do not require traditional, three-dimensional motion capture laboratories or complex wearable devices like inertial measurement units.

Video-based pose estimation is an emerging technology that applies computer vision (a subfield of artificial intelligence) to identify and track body positions automatically from digital videos (Cao, Hidalgo, Simon, Wei, & Sheikh, 2021; Mathis et al., 2018; Toshev & Christian Szegedy, 2014). Rather than using a set of specialized cameras or inertial measurement units, pose estimation requires only a common household video recording device such as a smartphone or tablet. Pose estimation has been used to conduct gait analysis to assess ostheoarthritis (Boswell et al., 2021), prostheses (Cimorelli, Patel, Karakostas, & Cotton, 2024), and chronic stroke (John et al., 2024; Lonini et al., 2022; Stenum, Hsu, Pantelyat, & Roemmich, 2024), among other clinical populations. Our group has developed and validated software to extract clinically relevant outcome measures (including gait parameters) using video-based pose estimation algorithms that are freely available. We have validated a variety of gait measurements that include walking speed, step time, step length, and lower limb joint angles against ground-truth motion capture across a variety of populations (Stenum, Rossi, & Roemmich, 2021) including individuals with stroke (John et al., 2024) or Parkinson’s disease (Stenum et al., 2024).

Here, we applied our video-based gait analysis software to measure spatiotemporal gait parameters from groups of non-frail, pre-frail, and frail older adults (as defined using the physical frailty phenotype). We hypothesized that our pose estimation-based software could be used to detect the following cross-sectional differences across frailty statuses: progressively slower walking speed, progressively shorter step lengths, and progressively longer step times with increasing frailty. Furthermore, we compared the pose estimation-based gait parameters against manually calculated parameters from the same videos and assessed the correlation of video-extracted gait parameters with clinical frailty scores, hypothesizing that the pose estimation-based parameters would correlate strongly with the manually calculated parameters and the clinical frailty scores.

## METHODS

We recruited 69 older adults (41 female, 28 male; age: 79.0±6.1 years (mean±standard deviation); height: 65.4±3.9 in; weight: 167.7±38.3 lbs) to participate in this study. Study participants were recruited from the “Registry of Older Adults Who May Be Willing to Participate in Research” (IRB# NA_00013162) on the Johns Hopkins Bayview Medical Campus. This registry is comprised of community-dwelling adults, aged 65 years or older, living in the Baltimore Metropolitan area. For each participant, we collected demographic information (Table 1) and physical frailty phenotype scores (Fried et al., 2001). Participants walked at both self-selected (SS) and fastest comfortable (FAST) paces along a 4.67 meter walkway. Participants performed trials, i.e. walking once back and forth along the walkway, two times at each pace for a total of four trials. As participants walked, we collected frontal plane video recordings (resolution: 1920 × 1080, frame rate: 30 Hz) with a Samsung Galaxy Tab A7 placed at the end of the walkway, 4.67 meters from the starting location (i.e., capturing a frontal view of the participant; Figure 1). For most participants, we tracked their gait as they walked along the walkway toward the camera. For participants using walkers, we instead tracked their gait as they walked away from the camera so that the walker would not obstruct the camera view of the body. *Video-based gait analysis*

**Figure 1.**
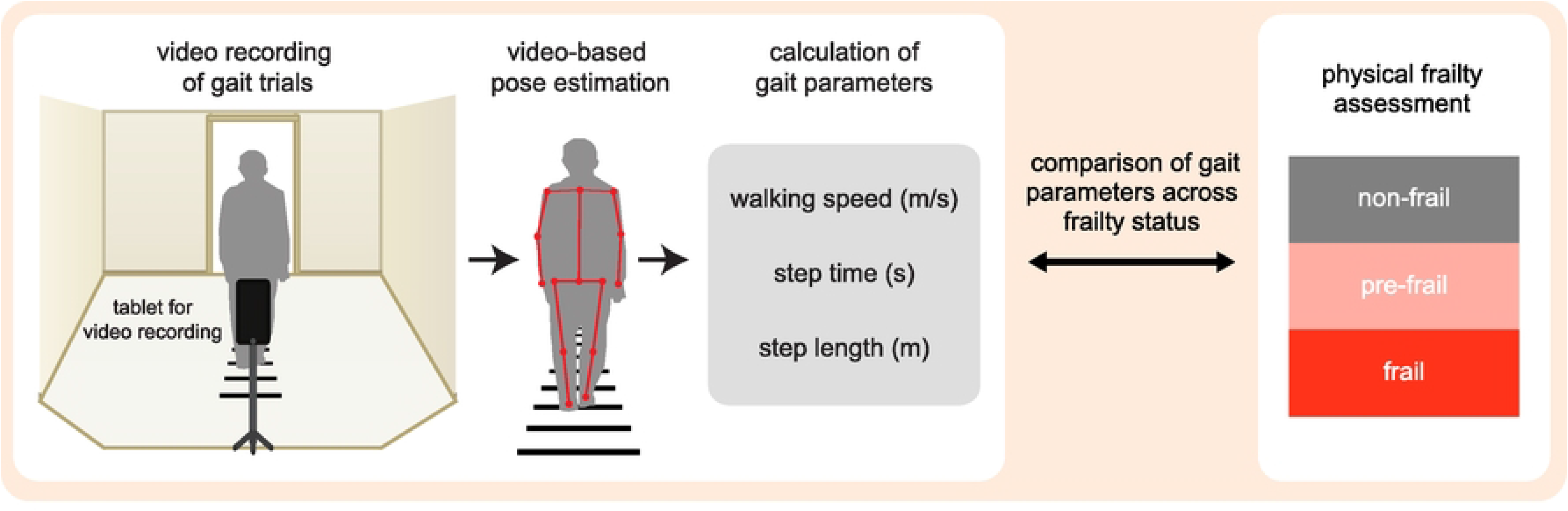
Schematic depicting the workflow from data collection through data analysis. Frontal-plane walking videos are recorded with a tablet mounted a tripod. Positions of body landmarks, i.e., keypoints, are extracted from videos using OpenPose pose estimation. Gait parameters are calculated from keypoints using a previously published video-based gait analysis pipeline. Gait parameters are compared across frailty statuses.

**Table 1.** Demographics of study participants.

| **Demographics** | **Description** | **Number** | **Percentage** |
| --- | --- | --- | --- |
| **Race** | Black or African American | 11 | 16.2 |
|  | White (Not Hispanic or Latino) | 54 | 79.4 |
|  | White (Hispanic or Latino) | 1 | 1.5 |
|  | Asian | 2 | 2.9 |
| **Gender** | Female | 40 | 58.8 |
|  | Male | 28 | 41.2 |
| **Age** | 60-69 | 6 | 8.8 |
|  | 70-79 | 31 | 45.6 |
|  | 80-89 | 31 | 45.6 |
| **Diagnosis** | Non-Frail | 25 | 36.8 |
|  | Pre-Frail 1 | 21 | 30.9 |
|  | Pre-Frail 2 | 4 | 5.9 |
|  | Frail 3 | 15 | 22.1 |
|  | Frail 4 | 3 | 4.4 |
|  | Frail 5 | 0 | 0.0 |

First, we used OpenPose – a freely available human pose estimation algorithm – to track body landmark positions from the videos (Cao, Simon, Wei, & Sheikh, 2017). The OpenPose BODY_25 model defines 25 two-dimensional keypoints representing body landmarks. We then input the keypoints into a previously established custom gait analysis pipeline implemented in MATLAB (Figure 1) (Stenum et al., 2024). This pipeline established a relationship between distance traversed (which, from the camera perspective, is depth change) and pixel size of the person by using two measured distances at fixed positions labelled with tape. This relationship then provided a spatial reference for measuring spatiotemporal gait parameters (e.g., walking speed, step length). We calculated step time by measuring the time from ipsilateral heel-strike to contralateral heel-strike. We first approximated heel-strikes automatically using our video-based gait analysis pipeline that defined heel-strikes using the vertical distance between left and right ankle keypoints where, when the participant walked toward the camera, positive peaks correspond to right heel-strikes and negative peaks correspond to left heel-strikes. To provide a ground-truth for validating the pose estimation-based step times, we also manually labelled heel-strikes and used them to calculate true step times as the time from ipsilateral heel-strike to contralateral heel-strike. Note that a ground-truth for spatial information (e.g., step lengths derived from three-dimensional motion capture data) was not available.

### Statistical analysis

To validate the pose estimation-based measurements of step time in our sample of older adults, we compared the pose estimation-based and manual step times using least-squares linear regression (analyzing step times during the SS and FAST trials separately). Specifically, we computed Pearson correlation coefficients and calculated Bland-Altman limits of agreement and corresponding margins of error (MoE) (Bland & Altman, 1986). Finally, we also computed mean absolute error (MAE) between the two methods.

We next compared the gait parameters across the different frailty statuses (non-frail, pre-frail, and frail) to assess potential cross-sectional differences. For each outcome measure (step time, step length, and walking speed) we used a two-way repeated measures ANOVA with pace (SS, FAST) as a within-subject factor and frailty status (non-frail, pre-frail, frail) as a between-subject factor. We also performed a one-way ANOVA to compare the changes in gait speed that the participants were able to achieve between the SS and FAST trials as a measure of gait reserve, or the capacity to increase walking speed beyond one’s preferred speed. Additionally, we tested the relationship between gait parameters and the binary gait slowness score from the physical frailty phenotype: walking speed, step time, and step length were tested using a two-way repeated measures ANOVA and change in walking speed was tested using a t-test. We report ANOVA results using the F-statistic, p-value, and effect size as measured by partial eta squared (*η*^2^). We applied Bonferonni-Holm corrections for multiple comparisons as post-hoc hypothesis tests where appropriate. We report these results using p-values and effect sizes, which were represented by Cohen’s d. For all tests, we used a significance level of *α*=0.05. We conducted linear regression and Bland-Altman analyses using MATLAB R2023b and all other statistical analyses using JASP 0.95.4.0.

## RESULTS

### Physical frailty phenotype scores

We excluded one participant from our study due to lack of usable video data, leaving 68 participants with data suitable for analysis. Participants exhibited a range of scores on the physical frailty phenotype: 25 participants were classified as non-frail (score=0), 25 participants were classified as pre-frail (score=1 or 2), and 18 participants were classified as frail, score≥3). *Validation of pose estimation-based step time measurements*

Pose estimation-based step time measurements closely agreed with manually identified step times (Figure 2). Step times from each measurement method were significantly correlated for both SS (R^2^=0.93, y=1.02x-0.03, p<0.001, MAE=0.025s) and FAST (R^2^=0.80, p<0.001, y=1.02x–0.01, MAE=0.034s) trials. Bland-Altman analysis of SS step times identified 95% limits of agreement of -0.082 to 0.052s (MoE: 0.014s). For FAST step times, the 95% limits of agreement were -0.114 to 0.110s (MoE: 0.023s).

**Figure 2.**
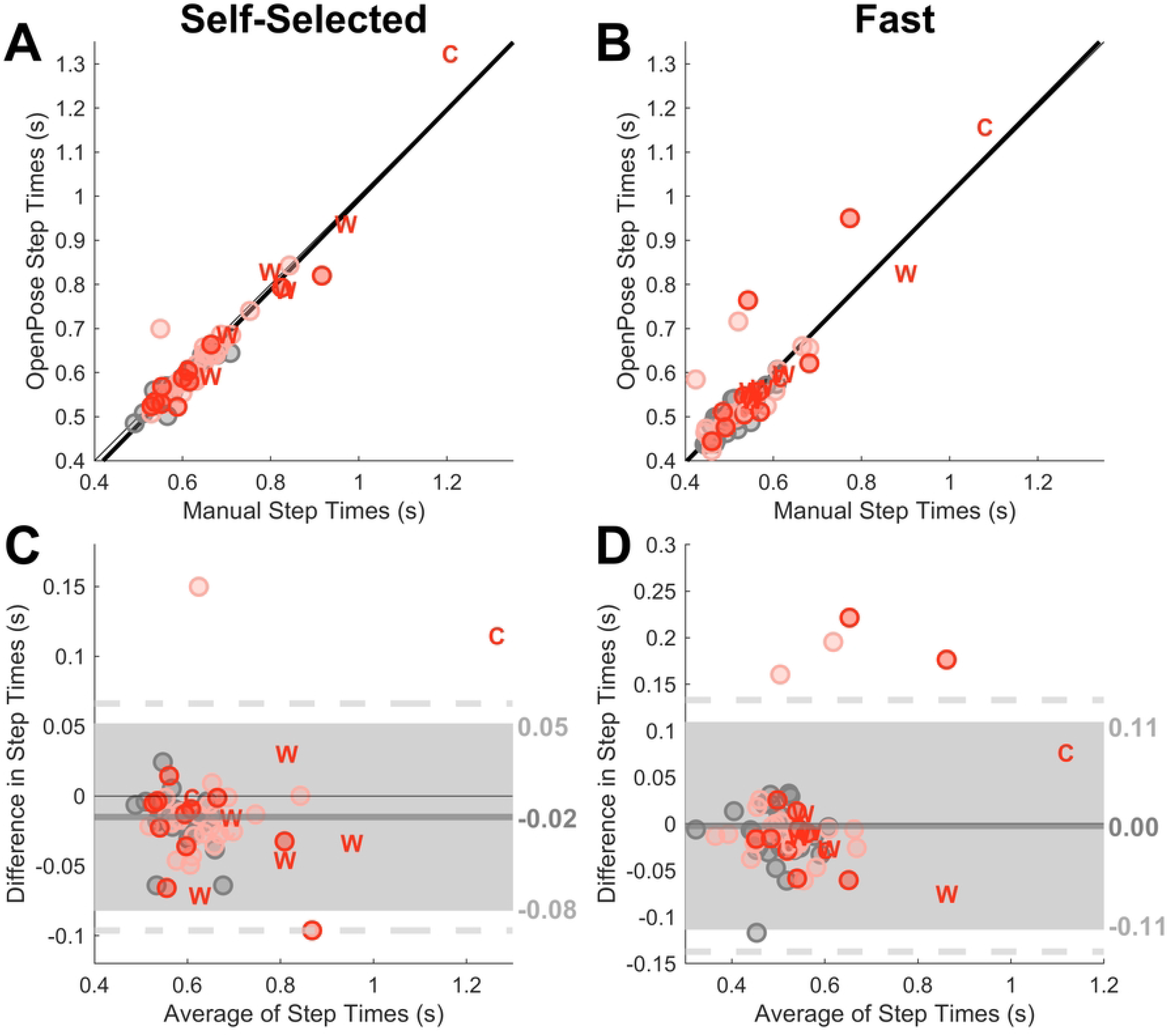
Correlation plots and Bland-Altman plots from the step time validation comparing step times calculated with OpenPose keypoint data against and manually identified step times. (A) Scatterplot of automatic and manual step times for SS walking. Circles depict data step times for individual participants, with grey circles indicating NF participants, pink circles indicating PF participants, and red circles indicating F participants. Data points from participants using walkers and canes are marked as “W” and “C”, respectively, and the text colors correspond with frailty status. The thick black lines represent a linear regression line fit to the dataset. The thin grey line depicts the line of equality (y=x). (B) Scatterplot of FAST step times in the same format as panel A. (C) Bland-Altman plot of SS step times, with average of step times on the x-axis and difference in step times (automatic-manual) on the y-axis. Circles represent individual participants. Canes and walkers are again labelled “C” and “W”. The thin black line depicts y=0, and the thick grey line depicts the mean of differences, which is labeled to the right of the graph. The shaded region depicts the upper and lower limits of agreement, which are labeled to the right of the graph. Dashed lines depict a 95% margin of error above the upper limit of agreement and below the lower limit of agreement. (D) Bland-Altman plot of FAST step times in the same format as panel C.

### Gait differences across clinical frailty statuses measured using video-based pose estimation

#### Gait speed

We observed significant main effects of both pace (F(1,65)=119.93, *η*^2^=0.65, p<0.01) and frailty status (F(2,65)=18.75, *η*^2^=0.37, p<0.01) on pose estimation-based measurements of walking speed. We also observed a significant pace × frailty status interaction (F(2,65)=3.25, *η*^2^=0.09, p=0.05). Post-hoc tests indicated a significant difference in walking speed between non-frail and pre-frail participants (p<0.01, d=0.79), between non-frail and frail participants (p<0.01, d=1.71), and between pre-frail and frail participants (p<0.01, d=0.92). Walking speeds were also significantly faster during FAST trials compared to SS trial s (p<0.01, d=1.14). During SS trials, walking speeds were 0.92±0.13 m/s in non-frail participants, 0.76±0.18 m/s in pre-frail participants, and 0.62±0.20 m/s in frail participants (Figure 3A). SS walking speed was significantly faster in non-frail participants compared to pre-frail participants (p<0.01, d=0.73), in non-frail participants compared to frail participants (p<0.01, d=1.39), and in pre-frail participants compared to frail participants (p<0.01, d=0.66). During FAST trials, walking speeds were 1.22±0.24 m/s in non-frail participants, 1.03±0.27 m/s in pre-frail participants, and 0.78±0.25 m/s in frail participants (Figure 3A). FAST walking speed was significantly faster in non-frail participants compared to pre-frail participants (p=0.01, d=0.85), in non-frail participants compared to frail participants (p<0.01, d=2.03), and in pre-frail participants compared to frail participants (p<0.01, d=1.19).

**Figure 3.**
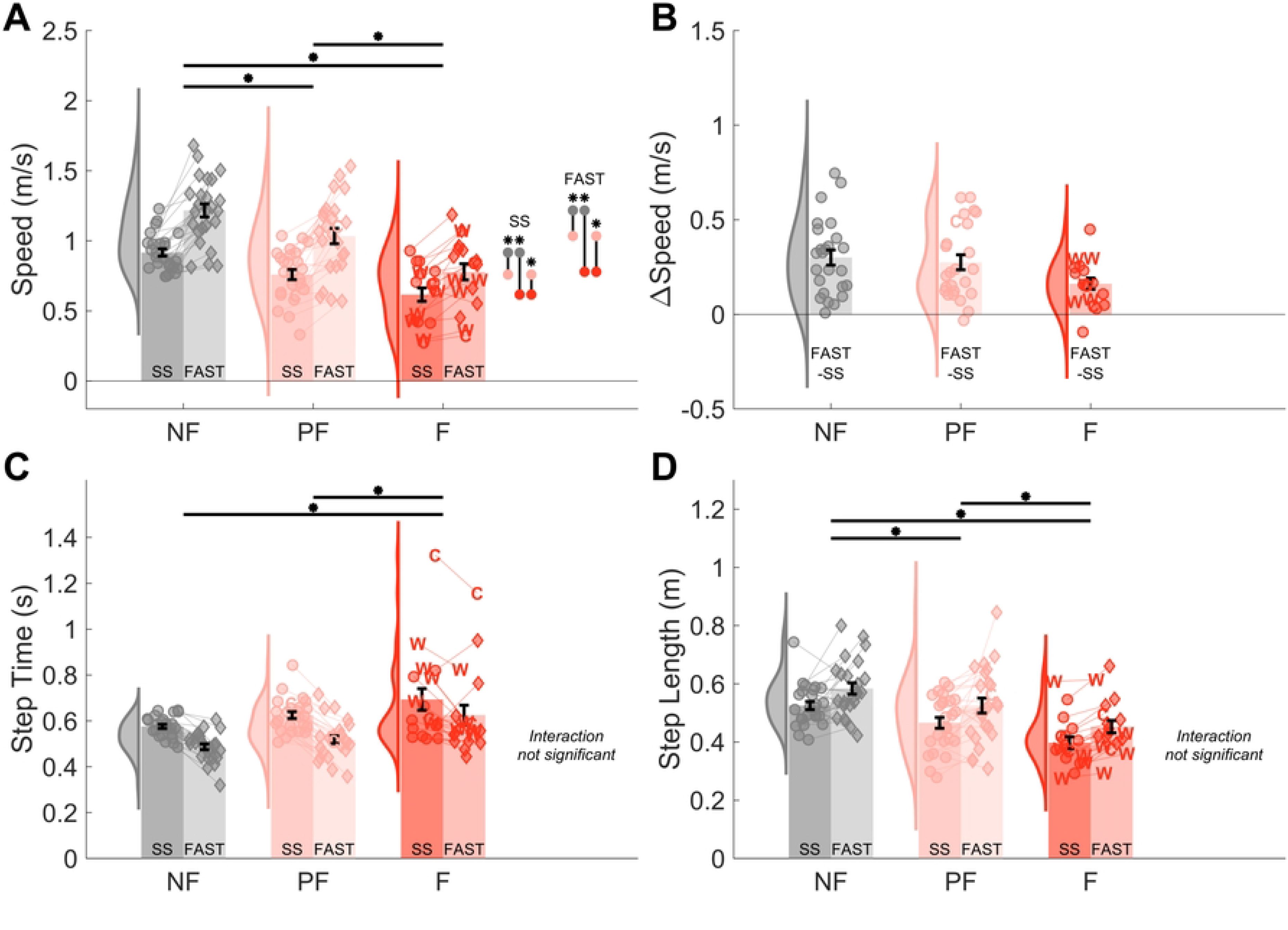
Gait parameters across frailty statuses. (A) Walking speed across frailty statuses. Frailty status (NF: grey, PF: pink, and F: red) is labeled on the x-axis walking speed is indicated on the y-axes. Pairs of like-colored bars indicate walking speeds for the corresponding frailty status broken down across self-selected (darker shade) and fast (lighter shade) paces. Black error bars indicate standard error of the mean. Individual participant average walking speed is indicated as circles for the self-selected pace and as diamonds for the fast pace. Light lines connect repeated measurements from the same participants across pace conditions. Distributions to the left of the bars indicate the kernel density estimate of the walking speed distribution pooled across self-selected and fast walking paces. Black lines and asterisks at the top of the plot indicate significant differences from post-hoc tests across the frailty status factor. Black lines with colored circles and asterisks to the right of the plot indicate significant differences from post-hoc tests on frailty status conditional on walking pace; circle colors correspond with frailty statuses. (B) Change in walking speed across frailty statuses. Format is consistent with panel (A), except that only a single bar representing change in speed from self-selected to fast is shown for each frailty status. (C-D) Step time and step length across frailty statuses. Format is the same as panel (A).

#### Change in gait speed from self-selected walking trials to fast walking trials

We observed a significant main effect of frailty status (F(2,65)=3.25, *η*^2^=0.09, p=0.05) on the change in gait speed measured between the SS and FAST trials. However, there was no significant change in gait speed between any pairwise frailty status comparisons following adjustment for multiple comparisons (Figure 3B).

#### Step time

We observed significant main effects of pace (F(1,65)=92.83, *η*^2^=0.59, p<0.01) and frailty status (F(2,65)=7.43, *η*^2^=0.19, p<0.01) on step time but no significant pace × frailty status interaction (F(2,65)=1.40, *η*^2^=0.04, p=0.25). Step times were significantly longer in frail participants compared to non-frail (p<0.01, d=1.12) and pre-frail (p=0.02, d=0.77) participants, but there was no significant difference in step times between pre-frail and non-frail (p=0.19, d=0.35). Step times were also significantly longer during SS trials compared to FAST trials (p<0.01, d=0.77). During SS trials, step times were 0.58±0.05 s in non-frail participants, 0.63±0.07 s in pre-frail participants, and 0.69±0.20 s in frail participants (Figure 3C). During FAST trials, step times were 0.49±0.06 s in non-frail participants, 0.52±0.08 s in pre-frail participants, and 0.63±0.18 s in frail participants (Figure 3C).

#### Step length

We observed significant main effects of pace (F(1,65)=27.58, *η*^2^=0.30, p<0.01) and frailty status (F(2,65)=11.96, *η*^2^=0.27, p<0.01) on step length but no significant pace × frailty status interaction (F(2,65)=0.02, *η*^2^=0.00, p=0.98). Step lengths were significantly longer in non-frail participants compared to pre-frail (p=0.02, d=0.61) and frail (p<0.01, d=1.34) participants and in pre-frail participants compared to frail participants (p=0.02, d=0.73). Step lengths were also significantly longer during FAST trials compared to SS trials (p<0.01, d=0.59). During SS trials, step lengths were 0.53±0.07 m in non-frail participants, 0.47±0.09 m in pre-frail participants, and 0.40±0.08 m in frail participants (Figure 3D). During FAST trials, step lengths were 0.59±0.10 m in non-frail participants, 0.53±0.13 m in pre-frail participants, and 0.45±0.09 m in frail participants (Figure 3D).

### Gait differences across older adults with and without clinically observed gait slowness

#### Gait speed

We observed significant main effects of pace (F(1,66)=45.53, *η*^2^=0.41, p<0.01) and slowness score (F(1,66)=34.45, *η*^2^=0.34, p<0.01) and a significant pace × slowness score interaction (F(1,66)=7.00, *η*^2^=0.10, p=0.01) on walking speed. Across slowness scores on the physical frailty phenotype, SS walking speeds were 0.83±0.18 m/s for participants with absence of gait slowness and 0.53±0.17 m/s for participants with presence of gait slowness (Figure 4A). Participants with gait slowness scores of 0 walked significantly faster than participants with scores of 1 (p<0.01, d=1.39) during the SS trials. Similarly, FAST walking speeds were 1.11±0.26 m/s for participants with gait slowness scores of 0 and were 0.64±0.20 m/s for participants with scores of 1 (Figure 4A). Participants with gait slowness scores of 0 also walked significantly faster than participants with scores of 1 (p<0.01, d=2.12) during the FAST trials.

**Figure 4.**
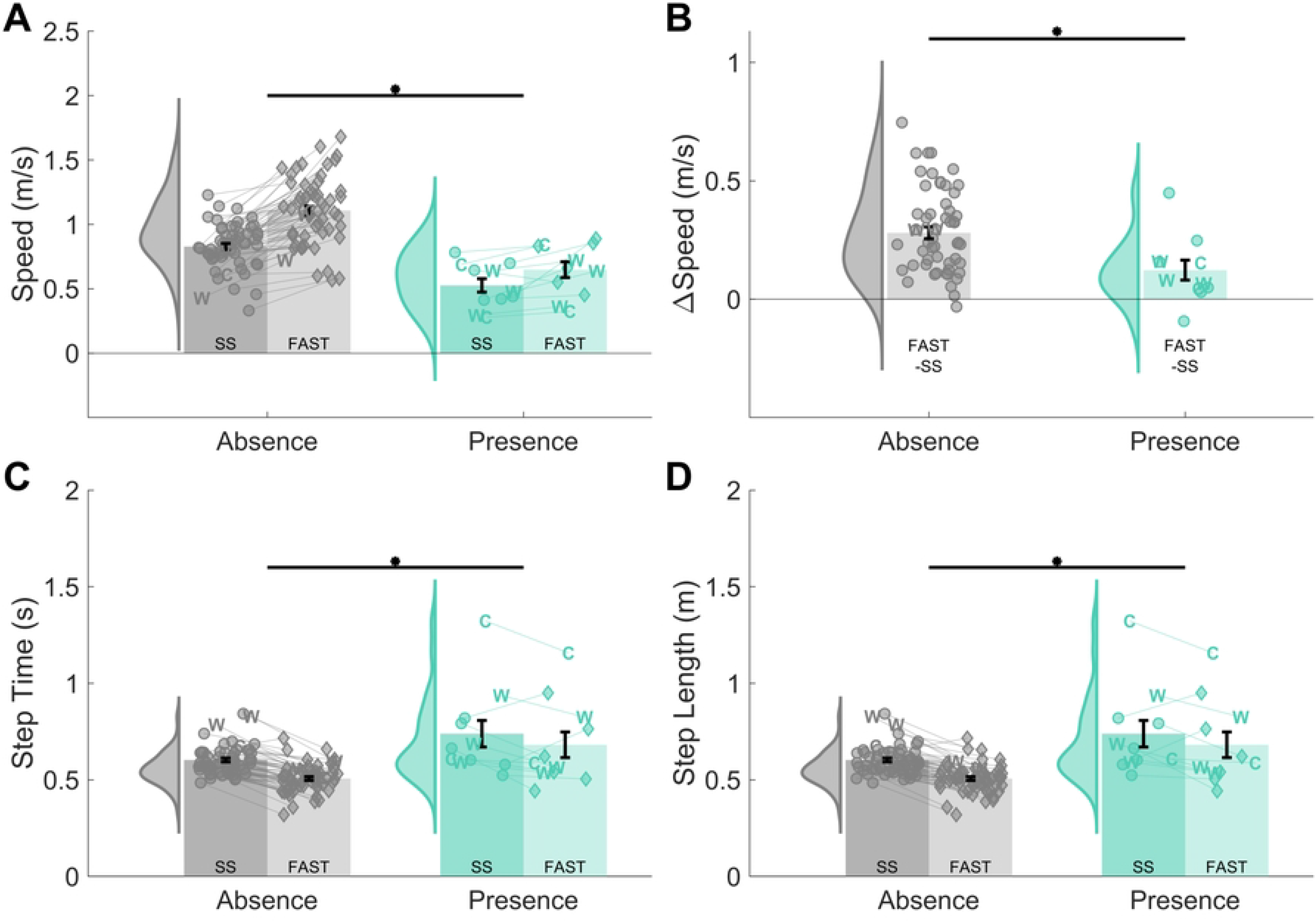
Gait parameters across slowness scores. (A) Walking speed across slowness score. Slowness score (absence of gait slowness: grey, presence of gait slowness: mint green) is labeled on the x-axis walking speed is indicated on the y-axes. Pairs of like-colored bars indicate walking speeds for the corresponding slowness score broken down across self-selected (darker shade) and fast (lighter shade) paces. Black error bars indicate standard error of the mean. Individual participant average walking speed is indicated as circles for the self-selected pace and as diamonds for the fast pace. Light lines connect repeated measurements from the same participants across walking pace conditions. Distributions to the left of the bars indicate the kernel density estimate of the walking speed distribution pooled across self-selected and fast walking paces. Black lines and asterisks at the top of the plot indicate significant differences from post-hoc tests across the slowness score factor. (B) Change in walking speed across slowness scores. Format is consistent with panel (A), except that only a single bar representing change in speed from self-selected to fast is shown for each slowness score. (C-D) Step time and step length across slowness scores. Format is the same as panel (A).

#### Change in gait speed from SS to FAST

We observed a statistically significant difference in gait speed change across slowness scores of 0 and 1 (p=0.01, d=0.34). Gait speed change was 0.28±0.19 m/s for slowness scores of 0 and 0.12±0.14 m/s for slowness scores of 1 (Figure 4B).

#### Step time

We observed significant main effects of pace (F(1,66)=39.80, *η*^2^=0.38, p<0.01) and slowness score (F(1,66)=20.56, *η*^2^=0.24, p<0.01) but no significant pace × frailty status interaction (F(1,66)=2.54, *η*^2^=0.04, p=0.12) on step time. Step times were significantly higher in participants with observed gait slowness than participants without observed gait slowness (p<0.01, d=1.41). Step times were also significantly higher during SS trials compared to FAST trials (p<0.01, d=0.70). Across gait slowness scores on the physical frailty phenotype, SS step times were 0.60±0.08 s for participants with absence of gait slowness and 0.74±0.23 s for participants with presence of gait slowness (Figure 4C). FAST step times were 0.51±0.07 s for participants with absence of gait slowness and were 0.68±0.22 s for participants with presence of gait slowness (Figure 4C).

#### Step length

We observed significant main effects of pace (F(1,66)=13.26, *η*^2^=0.17, p<0.01) and slowness score (F(1,66)=25.39, *η*^2^=0.28, p<0.01) but no significant pace × frailty status interaction (F(1,66)=0.24, *η*^2^=0.00, p=0.63) on step length. Step lengths were significantly higher in participants without observed gait slowness than participants with observed gait slowness (p<0.01, d=1.47). Step lengths were also significantly higher during FAST trials compared to SS trials (p<0.01, d=0.56). Across slowness scores on the physical frailty phenotype, SS step lengths were 0.49±0.09 m for participants with absence of gait slowness and 0.36±0.05 m for participants with presence of gait slowness (Figure 4D). Similarly, FAST step lengths were 0.55±0.11 m for participants with absence of slowness and were 0.40±0.05 m for participants with presence of gait slowness (Figure 4D).

#### Accounting for non-normality and potential age effects

We noted that there were instances of non-normality largely driven by outliers within some of our analyses. Non-normality could affect results from these statistical tests, so we re-performed relevant analyses with steps taken to address non-normality (Supplemental Document 1). This follow-up analysis yielded similar results to our initial analysis and did not change the primary conclusions of this study. We also conducted a follow-up analysis to test the effect of age on our results (Supplemental Document 1). This analysis did not identify a significant relationship between age and any gait parameter. Consequently, we were satisfied that our initial analysis, which did not account for age as a factor, was appropriate.

## DISCUSSION

Here, we applied our custom video-based gait analysis software to collect walking data on non-frail, pre-frail, and frail older adults. First, we validated video-based step time measurements against manually identified step time measurements. This validation showed that automated video-based measurements closely agreed with manual measurements. Next, we compared gait parameters (walking speed, change in walking speed, step time, and step length) across adults with different frailty statuses walking at self-selected and fastest comfortable paces. This analysis identified a significant effect of both frailty status and pace on all tested gait parameters. We also compared gait parameters across clinically assessed gait slowness scores at both paces and similarly found statistically significant effects of slowness score on all gait parameters.

The results of the frailty status analysis show the potential to measure and track features of frailty using simple video-based gait measures. In terms of effect size, walking speed exhibited the most distinct differences across frailty status (non-frail vs. frail at self-selected speed: d=1.39), but step length (non-frail vs. frail at self-selected speed: d=1.32) and step time (non-frail vs. frail at self-selected speed: d=1.03) still exhibited strong effect sizes. These results are consistent with prior studies which have shown that, among gait metrics, walking speed is the strongest indicator of frailty with effect sizes of 0.76–6.17 (Schwenk et al., 2013).

We also note that progressing from the self-selected to fast walking paces increased effect size for walking speed (significant pace × frailty status interaction (F(2,65)=3.25, *η*^2^=0.09, p=0.045; non-frail vs. frail at FAST pace: d=2.03 compared to SS pace: d = 1.39). Prior work suggested that fast walking might provide better insight into frailty-related mobility impairment than self-selected walking given that walking at a faster pace requires greater physiological reserve (Schwenk et al., 2013). Indeed, fast walking is more sensitive than self-selected walking in detecting aging-related gait changes (Ko, Hausdorff, & Ferrucci, 2010) or cognitive decline (Deshpande, Metter, Bandinelli, Guralnik, & Ferrucci, 2009; Fitzpatrick et al., 2007).

However, it is not yet clear if fast walking is a more sensitive test for identifying frailty status. Prior studies have not demonstrated conclusive differences between self-selected and fast walking pace effect sizes (Middleton et al., 2016; Montero-Odasso et al., 2011), but one study identified increased effect sizes for variability metrics under the fast condition (Montero-Odasso et al., 2011). Our study provides further evidence in support of fast walking as a clearer differentiator of frailty status than self-selected walking.

Despite this increased statistical effect size of fast compared to self-selected walking paces, we note that change in walking speed (i.e., walking speed reserve) did not differ significantly among frailty statuses (with an effect size of only *η*^2^=0.09). This walking speed reserve metric has been theorized as a potential indicator of frailty, but, consistent with prior studies (A Middleton et al., 2016), our results do not indicate a strong relationship between walking speed reserve and frailty, whereas raw gait speed at both self-selected and fastest comfortable pace exhibited a strong relationship to frailty status.

The gait slowness score analysis provided confirmation that video-based gait measures are consistent with clinically observed gait slowness as defined by the physical phenotype. As expected, walking speed and all other tested gait parameters differed significantly between participants with clinically observed gait slowness on the physical phenotype. Notably, there is overlap between the walking speed distributions of participants with and without gait slowness. This is likely because, under the physical frailty phenotype, the walking speed threshold for gait slowness is adjusted for each patient according to the bottom 20^th^ percentile for adults of the same height and sex. The same method could likewise be applied to our video-based walking speed measurements.

The results of this study have potential value for clinical frailty diagnosis. Currently, frailty classification relies on composite clinical assessments like the physical frailty phenotype, but an automated gait assessment could streamline assessments and provide quantitative movement data to track functional status and frailty over time. The broad accessibility of video-recording devices such as smartphones or tablets suggest that this scalable approach could meet both the practical and technical criteria for broader clinical implementation. Further development of this approach could facilitate faster and more reliable means of identifying frailty in older adults.

We acknowledge some limitations to our study. In some instances, participants used canes or walkers to support their walking, and OpenPose occasionally detects keypoints on these devices instead of on the participant. We adapted our data processing methods and used an alternative workflow for participants with walkers to track their gait as they walked away from rather than towards the camera. Also, the walkway used for our analysis was relatively short at roughly four meters, which only allowed us to record a few strides of continuous walking at a time; consequently, we recorded a relatively small sample of gait cycles. However, the use of a small walkway is also a strength of this study since it represents the likely constraints on available walking space in clinical evaluation rooms, and a four-meter walkway length was recommended by the International Academy on Nutrition and Aging Task Force (Abellan Van Kan et al., 2009). Future work on this project will expand this analysis to include more gait parameters such as step width and measures of gait variability. We will also compare gait measures and frailty status longitudinally within this cohort to evaluate the potential of video-based gait measurements to predict immediate and future frailty status.

In conclusion, this study demonstrated that a video-based measurement system can measure differences in spatiotemporal gait parameters across frailty statuses in older adults. Furthermore, a validation of step time measurements demonstrated the close agreement between step times detected using our gait analysis pipeline and manually annotated step times. These results show that video-based gait analysis methods could provide a new, scalable approach for measuring and tracking frailty status in older adults.

## DATA AVAILABILITY

The dataset from this study is publicly available at this Github repository: https://github.com/KalebEBurch/PoseEstimation_Frailty_GaitAnalysis

## ACKNOWLEDGEMENT

This work was supported by the Johns Hopkins University Claude D. Pepper Older Americans Independence Center, funded by the National Institute on Aging and National Institutes of Health (grant P30AG021334 to PMA); the Translational Aging Research Training Program (NIA Grant T32AG058527 to PMA); and the Precise Center, funded by the NIH (grant P50HD118624 to RTR).

